# SARS-CoV-2 Infection Breakthrough among the non-vaccinated and vaccinated: a Real World Evidence study based on Big Data

**DOI:** 10.1101/2022.02.22.21266830

**Authors:** A Perrella, M Bisogno, D’Argenzio, U Trama, E Coscioni, V Orlando, COVID CaRe group

**Affiliations:** Regional Task Force COVID-19, Campania Region, Naples, Italy; Directorate-General for Health Protection, Campania Region, Naples, Italy; Sinfonia Regional Health Information System of Campania Region, Naples, Italy; Division of Cardiac Surgery, AOU San Giovanni di Dio e Ruggi d’Aragona, Salerno, Italy; CIRFF (Center of Drug Utilization and Pharmacoeconomics), Department of Pharmacy, University of Naples Federico II, Naples, Italy; Hospital Health Direction, Infectious Disease Unit, Hospital A. Cardarelli, Naples, Italy; Regional crisis Unit COVID-19, Campania Region, Naples, Italy

**Keywords:** COVID-19, vaccination coverage, Real World Evidence study, Big Data, Machine Learning, SARS-COV-2, Vaccine

## Abstract

**Background:** SARS-CoV-2 infection after vaccination can occur because COVID-19 vaccines do not offer 100% protection. The aim of this study was to assess vaccination coverage among people nasopharyngeal swabs, disease symptoms and type of hospitalisation (Intensive Care Unit) between the non-vaccinated and the effective dose vaccinated and to evaluate vaccination trend over time.

**Methods:** A retrospective cohort study was carried out among people tested positive for COVID-19 in Campania Region using collected information from Health Information System of Campania Region (Sinfonia).

The status of vaccination was assess according to the following timetable: “non-vaccinated”; “Ineffective dose” vaccination; “Effective dose” vaccination.

Univariate and multivariate logistic regression models were conducted to evaluate the association between Intensive Care Unit (ICU) to COVID-19 and gender, age groups and vaccine.

To determine vaccine coverage in subjects who received an effective dose, trend changes over time were investigated using segmented linear regression models and breakpoints estimations.

Vaccination coverage was assessed by analysing the trend in the percentage of covid 19 positive subjects in the 9 months after vaccination with an effective dose stratified by age group and type of vaccine. Statistical analyses were performed using R platform

**Results:** A significant association with the risk of hospitalisation in Intensive Care Unit was the vaccination status of the subjects: subjects with ineffective dose (adjusted OR: 3.68) and subjects no-vaccination (adjusted OR: 7.14) were at three- and seven-times higher risk of hospitalisation in Intensive Care Unit, respectively, than subjects with an effective dose.

Regarding subjects with an effective dose of vaccine, the vaccine’s ability to protect against infection in the months following vaccination decreased.

The first breakpoints is evident five months after vaccination (β =1.441, p<0.001). This increase was most evident after the seventh month after vaccination (β =3.110, p<0.001).

**Conclusions:** COVID19 vaccines protect from symptomatic infection by significantly reducing the risk of ICU hospitalization for severe disease. However, it seems they have trend to decrease their fully protection against SARS-COV-2 after five months regardless age, sex or type of vaccine. Therefore it seems clear that those not undergoing vaccine had higher risk to develop clinically significant disease and being at risk of ICU stay. Thus, considering highest percentage of asymptomatic patients and that few data about their capacity to transmit SARS-CoV-2, third dose vaccination should be introduced as soon as possible while awaiting antivirals.Finally, a surveillance approach based on the use of integrated BIG Data system to match all clinical conditions too, offer a precise and real analysis with low incidence of errors in the categorization of subjects.

## Introduction

Since WHO declared the emergence of coronavirus disease 2019 (COVID-19) pandemic on March 11, 2020, over 5 million people have died worldwide, including over 130,000 people in Italy.^1^

Due to severe acute respiratory syndrome coronavirus-2 (SARS-CoV-2) infection impact on health system of all countries, some countries or pharmaceutical companies have promoted research protocol to find a cure or to develop a vaccine against the SARS-CoV-2.^2-4^

Currently several vaccines have been produced and authorized, being based on different technologies. In Italy at the end of 2020 and throughout 2021, following European Medical Agency (EMA), Italian Medicines and Healthcare products Regulatory Agency (AIFA) authorized the BNT162b2 mRNA (Pfizer-BioNTech) and ChAdOx1 nCoV-19 adenoviral (Oxford-AstraZeneca), CX-024414 mRNA (Moderna) and Ad26.COV2-S Adenoviral (J&J). Since the authorization of vaccine, thanks to extraordinary effort of all Italian healthcare workers and sense of civil responsibility of Italian population a high percentage of people were vaccinated and COVID19 contagious progressively decrease over the entire country during the first 8 months of 2021.^5^ Despite recent studies ^6-8^ there is still a lack of studies on vaccine efficacy based on real world data. The aim of this study was to assess vaccination coverage among people nasopharyngeal swabs, disease symptoms and type of hospitalisation (Intensive Care Unit) between the non-vaccinated and the effective dose vaccinated and to evaluate vaccination trend over time.

## Materials and Methods

### Data sources: Sinfonia

Sinfonia includes records on patient demographics and for ∼ 6 million residents, comprising a well-defined population in Italy (∼ 10% of the population of Italy).

Sinfonia collects information, encrypted and anonymized from Local Health Unit (LHU) whose are legal owner of the original data, in accordance with the privacy laws. All analyses on the data are therefore carried out on encrypted and anonymized data using transparent data encryption protocols.

It is complete and involves data management system that has been validated in previous studies. ^9-12^ During the pandemic emergency, the Regional Health Information System of Campania Region (Sinfonia) was implemented with all records related to COVID-19 in order to create a tool to support health governance in managing the COVID-19 emergency.

The aims of Sinfonia tool, based on previous experiences too^13^, were:

1. Applying data science methods to big data in order to assess pandemic trends
2. Creation of predictive algorithms through AI methods
3. ML analysis, performed according to the python scripting model (Spyder IDE 64bit ver), to perform predictive analysis on contagiousness.

The characteristics of Sinfonia are described in Supplementary Material 1

### Study design and cohort selection

A retrospective cohort study was carried out among people tested positive for COVID-19 in Campania Region since March 8, 2021, until 31 October 2021 using collected information from Health Information System of Campania Region (Sinfonia).

Nasopharyngeal swabs were collected by trained personnel of Regional Healthcare system and/or authorized and trained territorial laboratory staff. RT-PCR testing was performed with the use of standardized RT-PCR machine from Coronavirus Network Laboratory (CoroNetLab), with four genes analysis RdRP, S and N genes specific to SARS-CoV-2, and the E gene with results expressed as the cycle threshold (Ct). A Ct value of less than 30, which indicated an increased viral load, was used to determine infectivity.^14,15^

Were considered as fully positive only results when all 4 genes found to be amplified by Rt-PCR while in all other case results were considered doubt and repeated. ^14,15^ Participant consent was given for the release of all SARS-CoV-2 PCR test results before or after vaccination. All positive participants were followed-up until negative PCR test.

For all individuals being positive at nasal swab clinical symptoms were collected according to Italian National Health Institute. Typical COVID-19 symptoms were fever, cough, or change or loss of taste or smell. Participants were recorded as having other symptoms if they reported any of the following: shortness of breath, sore throat, runny nose, headache, muscle aches, extreme fatigue, diarrhoea, nausea or vomiting, or small itchy red patches on fingers or toes, on the follow-up questionnaire with a symptom onset date within 14 days before or after the PCR positive sample date.

Data extraction was conducted from Sinfonia every month to have e regular report of vaccine/positive trend and for final analysis on October 31, 2021. All collected data after ML algorithm were anonymized and encrypted according to transparent data encryption.

Briefly, AI based on ML algorithm, was used in data mining on Sinfonia to daily match records from SARS-COV-2 RT-PCR nasal swab and status of vaccination according to the following timetable:

i. Positive without vaccinated being considered “*non-vaccinated”*
ii. positive after 1^st^ dose vaccine (<15 or >15 days) or after two dose vaccine < 15 days and being considered “*Ineffective dose”* vaccination,
iii. positive after two dose with more than 15 days since second dose of BNT162b2 mRNA (Pfizer-BioNTech), ChAdOx1 nCoV-19 adenoviral (AstraZeneca) and CX-024414 mRNA (Moderna) or in case of Ad26.COV2-S Adenoviral (J&J) 60 days after one shot and being considered “E*ffective dose”* vaccination.

Flow chart is illustrated in Figure 1.

**Figure 1.**
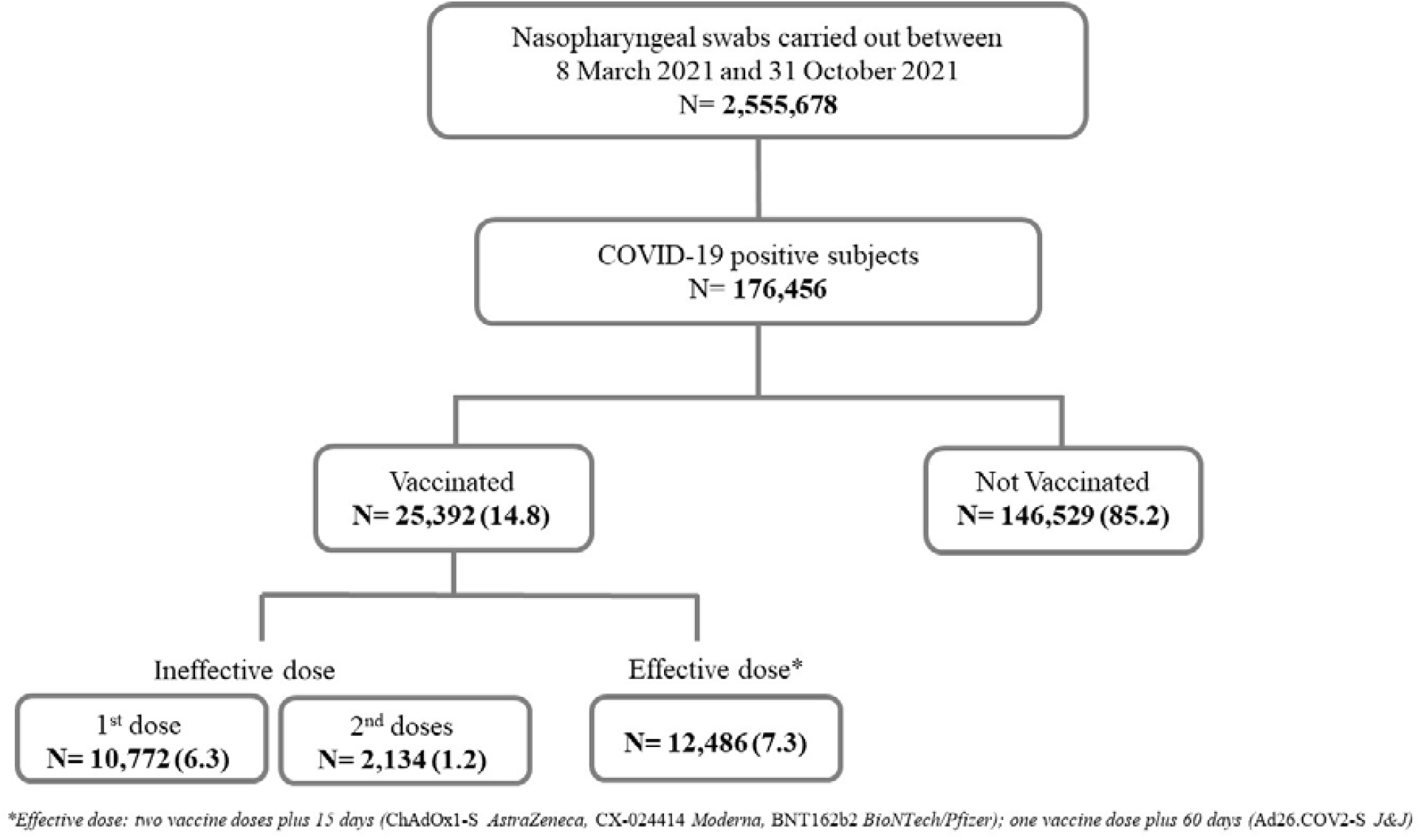
Flowchart: cohort selection

All subjects provided written informed consent to vaccination and data storage on a Big Data system management to collect all COVID-19 patients’ data and related clinical history (Symptoms, hospital admission and related follow-up, previous clinical status) according to European Privacy Policy to manage pandemic.

Time elapsed from second dose and onset of COVID-19 was calculated for all individuals to evaluate risk of infection in time dependent way. Further, once recognized a positive subject among those vaccinated was evaluated according to days elapsed since vaccine.

### Outcomes

The primary outcome was to assess the risk of intensive care unit (ICU) admission for COVID-19 between the non-vaccinated and the effective dose vaccinated.

Secondary outcome was to evaluate vaccination coverage, over time, stratified by age group and vaccine type

### Statistical analysis

The study population baseline characteristics were analyzed using descriptive statistics. Quantitative variables were described as counts and percentages. The chi-square test and t-test were performed to determine the difference between non-vaccinated and vaccinated subjects who tested positive for COVID-19. In particular, the vaccinated subjects were distinguished into two groups: vaccinated with an ineffective dose (one vaccine dose or two vaccine doses) and vaccinated with an effective dose (two vaccine doses plus 15 days: BNT162b2 mRNA (Pfizer-BioNTech), ChAdOx1 nCoV-19 adenoviral (AstraZeneca) and CX-024414 mRNA (Moderna); one vaccine dose plus 60 days: Ad26.COV2-S Adenoviral (J&J)).

Univariate and multivariate logistic regression models were conducted to evaluate the association between Intensive Care Unit (ICU) to COVID-19 and gender, age groups (ie, 40 - 59 years, 60 - 79 years, ≥ 80 years vs 0-39 years) and vaccine (Non-vaccination, Ineffective dose vs Effective dose). To determine vaccine coverage in subjects who received an effective dose, trend changes over time were investigated using segmented linear regression models and breakpoints estimations.

Breakpoints were identified testing differences in slope and intercepts of the trend and then different linear models were implemented. Changes in the slope segment indicated an impact of vaccination coverage on protection against COVID-19 infection.

Every linear model was expressed as follows: yt = a +b * t + et, where a was the intercept, b the slope and et the error term. Coefficients (β) were considered statistically significant with a P value < 0.05. The 95% confidence intervals (CIs) for each breakpoint were also obtained.

In addition, vaccination coverage was assessed by analysing the trend in the percentage of covid 19 positive subjects in the 9 months after vaccination with an effective dose stratified by age group and type of vaccine. Statistical analyses were performed using R platform (version 3.6, The R Formulation for Statistical Computing, Vienna, Austria).

## Results

During analysed period 8 March 2021 to 31 October 2021, in Campania Region, 2,555,678 nasal swabs were performed in subjects aged 18-98 years. Are showed in figure 1 of the total cohort of COVID-19 positive subjects, 85.2% were non-vaccinated (N= 146.529) and 14.8% (N= 25.392) were vaccinated. Of the 25,392 subjects who received at least one dose of vaccine, 7.5% (N= 12,906) received an ineffective dose; in comparison 7.3% (N=12,486) received an effective dose. Of the total 171,921 COVID-19 positive subjects, 51.2% were females.

The analysis stratified by age group showed that among the total cohort of COVID-19 positive subjects, 50.9% were aged 0 -39 years, 29.4% 40 - 59 years, 16.3% 60 - 79 years and 3.4% were aged more than 80 years.

Information of disease symptoms was not available for 34,119 subjects (19.8%) included in the analysis.

In particular, the percentage of subjects with severe and critical COVID-19 decreased in the cohort of vaccinated subjects compared to non-vaccinated subjects. Among a total of 482 subjects with severe symptoms 89.4% of the subjects are non-vaccinated, 7.5% of the subjects are vaccinated with a non-effective dose and 3.1% of the subjects are vaccinated with an effective dose.

Similarly, out of 57 subjects with critical symptoms 82.5% of the subjects are non-vaccinated, 10.5% of the subjects are vaccinated with a non-effective dose and 7.0% of the subjects are vaccinated with an effective dose.

Overall, 2.7% of the COVID-19 positive subjects were hospitalised and 0.1% were in intensive care unit.

Among hospitalised subjects the majority (83.7%) were non-vaccinated, 10.3% received a non-effective dose and 6.0%) received an effective dose.

Among subjects in Intensive Care Unit (ICU) the majority (90.5%) were non-vaccinated, 7.6% received a non-effective dose and 1.9% received an effective dose (Table 1).

**Table 1.** General characteristics of COVID-19 positive patients

|  | Total N (%) | No | Vaccination N (%) |  | Effective dose* |
| --- | --- | --- | --- | --- | --- |
|  |  |  | Ineffective dose |  |  |
|  |  |  | 1<sup>st</sup> dose | 2<sup>nd</sup> doses |  |
|  | 171,921 (100) | 146,529 (85.2) | 10,772 (6.3) | 2,134 (1.2) | 12,486 (7.3) |
| Gender |  |  |  |  |  |
| Male | 83,924 (48.8) | 71,693 (85.4) | 5,249 (6.3) | 1,168 (1.4) | 5,814 (6.9) |
| Female | 87,997 (51.2) | 74,836 (85.0) | 5,523 (6.3) | 966 (1.1) | 6,672 (7.6) |
| Age Groups |  |  |  |  |  |
| 0 - 39 years | 87,464 (50.9) | 79,686 (91.1) | 3,101 (3.5) | 1,099 (1.3) | 3,578 (4.1) |
| 40 - 59 years | 50,539 (29.4) | 42,042 (83.2) | 3,419 (6.8) | 462 (0.9) | 4,616 (9.1) |
| 60 - 79 years | 28,079 (16.3) | 21,331 (76.0) | 3,371 (12.0) | 372 (1.3) | 3,005 (10.7) |
| ≥ 80 years | 5,839 (3.4) | 3,470 (59.4) | 881 (15.1) | 201 (3.4) | 1,287 (22.0) |
| Vaccine type |  |  |  |  |  |
| ChAdOx1-S (AstraZeneca) | 6,067 (3.5) | - | 3,913 (64.5) | 190 (3.1) | 1,964 (32.4) |
| Ad26.COV2-S (J&J) | 1,181 (0.7) | - | 824 (69.8) | - | 357 (30.2) |
| CX-024414 (Moderna) | 1,774 (1.0) | - | 898 (50.6) | 91 (5.1) | 785 (44.3) |
| BNT162b2 (Pfizer-BioNTech) | 15,657 (9.1) | - | 5,422 (34.6) | 998 (6.4) | 9,237 (59.0) |
| Disease Symptoms |  |  |  |  |  |
| Asymptomatic | 112,251 (65.3) | 95,482 (85.1) | 7,343 (6.5) | 1,526 (1.4) | 7,900 (7.0) |
| Paucysinintomatic | 14,339 (8.3) | 12,542 (87.5) | 865 (6.0) | 134 (0.9) | 798 (5.6) |
| Mild | 10,673 (6.2) | 9,368 (87.8) | 599 (5.6) | 60 (0.6) | 646 (6.1) |
| Severe | 482 (0.3) | 431 (89.4) | 35 (7.3) | 1 (0.2) | 15 (3.1) |
| Critical | 57 (0.03) | 47 (82.5) | 6 (10.5) | - | 4 (7.0) |
| Not available | 34,119 (19.8) | 28,659 (84.0) | 1,924 (5.6) | 413 (1.2) | 3,123 (9.2) |
| Hospitalisation |  |  |  |  |  |
| Yes | 4705 (2.7) | 3,939 (83.7) | 431 (9.2) | 52 (1.1) | 283 (6.0) |
| No | 167,216 (97.3) | 142,590 (85.3) | 10,341 (6.2) | 2,082 (1.2) | 12,203 (7.3) |
| Intensive Care Unit (ICU) |  |  |  |  |  |
| Yes | 211 (0.1) | 191 (90.5) | 16 (7.6) | - | 4 (1.9) |
| No | 171,710 (99.9) | 146,338 (85.2) | 10,756 (6.3) | 2,134 (1.2) | 12,482 (7.3) |

\*Effective dose: two vaccine doses plus 15 days (ChAdOx1-S, CX-024414, BNT162b2); one vaccine dose plus 60 days (Ad26.COV2-S)
#Vaccine type: only for subjects who have received at least one dose of vaccine

Table 2 reports the results of the univariate and multivariate logistic regression analyses, which showed that three independent variables made a statistically significant contribution to the model: gender, age, and vaccination status of the subjects were the main determinants of the risk of hospitalisation in Intensive Care Unit (ICU) to COVID-19.

**Table 2.** Univariate and multivariate logistics regression of the risk of hospitalisation in Intensive Care Unit (ICU) to COVID-19

|  | Unadjusted<br>OR (95% CI) | p-Value | Adjusted<br>OR (95% CI) | p-Value |
| --- | --- | --- | --- | --- |
| **Gender** |  |  |  |  |
| Male (vs *Female*) | 1.59 (1.20 - 2.09) | 0.001* | 1.70 (1.29 - 2.24) | <0.001* |
| **Age groups** |  |  |  |  |
| 40 - 59 years (vs 0–39 years) | 5.57 (3.05 - 10.14) | <0.001* | 5.99 (3.28 - 10.90) | <0.001* |
| 60 - 79 years (vs 0–39 years) | 29.73 (17.13 - 51.57) | <0.001* | 33.53 (19.31 - 58.23) | <0.001* |
| ≥ 80 years (vs 0–39 years) | 20.39 (10.21 - 40.69) | <0.001* | 29.04 (14.48 - 58.27) | <0.001* |
| **Vaccination** |  |  |  |  |
| Ineffective dose (vs *Effective dose*) | 3.94 (1.31 - 11.79) | 0.014* | 3.68 (1.23 - 11.02) | 0.020* |
| No-vaccination (vs *Effective dose*) | 4.11 (1.52 - 11.06) | 0.005* | 7.14 (2.64 - 19.27) | <0.001* |

**Abbreviations:** CI, confidence interval; OR, odds ratio
\**p-value* <0.05 was considered to be statistically significant.

A significant association with the risk of hospitalisation in Intensive Care Unit was the gender. Males (adjusted odds ratio [OR]: 1.70; 95% CI: 1.29 - 2.24, p value <0.001) were at almost two times higher risk of hospitalization in Intensive Care Unit than females. Similarly, a strong significant association with the risk of hospitalisation in Intensive Care Unit was the age. Subjects aged 60-79 years (adjusted odds ratio [OR]: 33.53; 95% CI: 19.31–58.23, p value <0.001) and subjects aged more than 80 years (adjusted odds ratio [OR]: 29.04; 95% CI: 14.48 - 58.27, p value <0.001) were more than thirty times and twenty-nine times, respectively, likely to the risk of hospitalization in Intensive Care Unit compared to subjects aged 0-39 years. Equivalent results were found for the vaccination status of the subjects: subjects with ineffective dose (adjusted OR: 3.68; 95% CI: 1.23 - 11.02, p value <0.001) and subjects no-vaccination (adjusted OR: 7.14; 95% CI: 2.64 - 19.27, p value <0.001) were at three- and seven-times higher risk of hospitalisation in Intensive Care Unit, respectively, than subjects with an effective dose.

In Campania region, from 8 March 2021 to 31 October 2021, 3.699.683 subjects received a complete vaccine schedule. Of all those vaccinated with effective dose 12,486 developed COVID-19, so the prevalence of COVID-19 positive vaccinated subjects was 0.33%.

Regarding 12,486 subjects with an effective dose of vaccine, the ability of the vaccine to protect against infection in the months following vaccination has been was investigated through the estimation of breakpoints, i.e., points in which data show deviations from stability in the background trend. Indeed, figure 2 shows the trend for the percentage of subjects testing positive for COVID-19 over the 9 months after an effective dose. Two breakpoints were identified from the analysis. The first breakpoints is evident five months after vaccination (β =1.441, p<0.001). This increase was most evident after the seventh month after vaccination (β =3.110, p<0.001).

**Figure 2.**
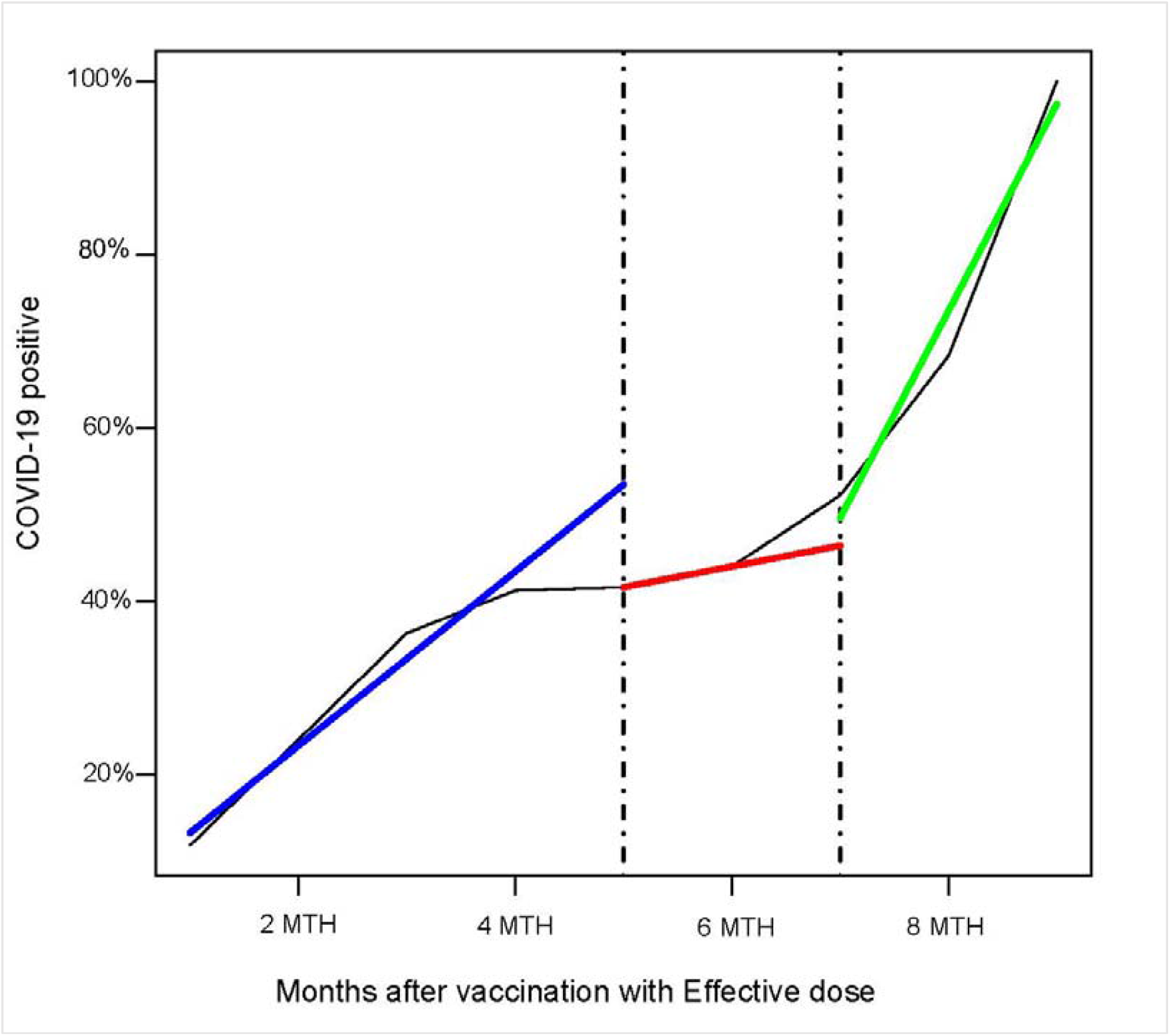
Segmented linear regression models

The analysis stratified by age group showed a similar trend in subjects aged 0-39 years, 40-59 years and 60-79 years in terms of increased number of COVID-19 positive patients (about 50%) up to the sixth month after vaccination. On the contrary, among subjects aged over 80 years, the trend up to the sixth month after vaccination was different: the percentage of positive subjects did not exceed 40%.

On the other hand, six months after vaccination, the trend was similar in all age groups (figure 3). The analysis stratified by vaccine type, however, showed that all subjects vaccinated with Ad26.COV2-S Adenoviral (J&J) (n=357) were positive for COVID-19 within one month after the effective dose (third month).

**Figure 3.**
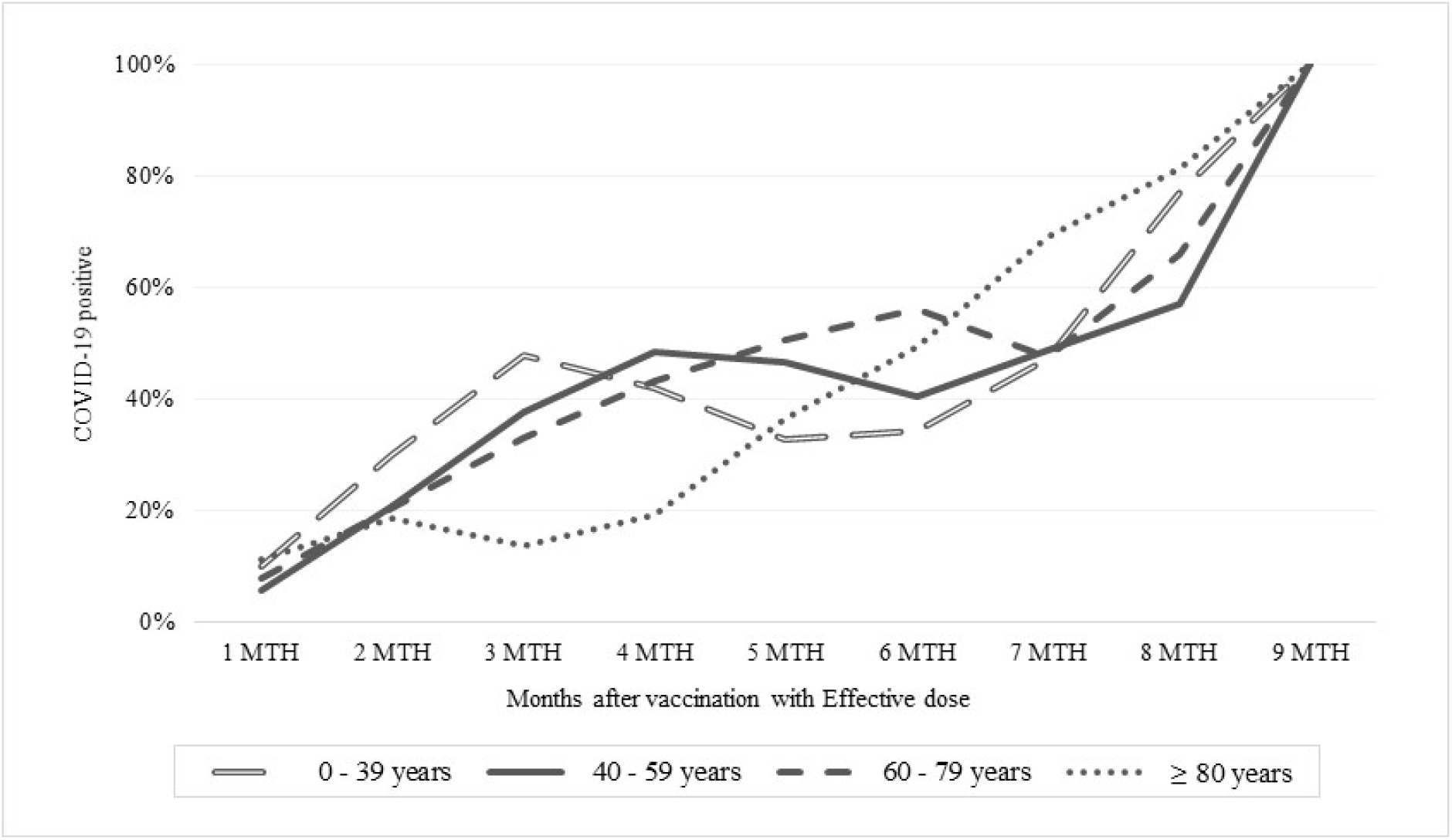
Percentage of COVID-19 positive patients stratified by number of months after vaccination with effective dose and stratified by age groups

**Figure 4.**
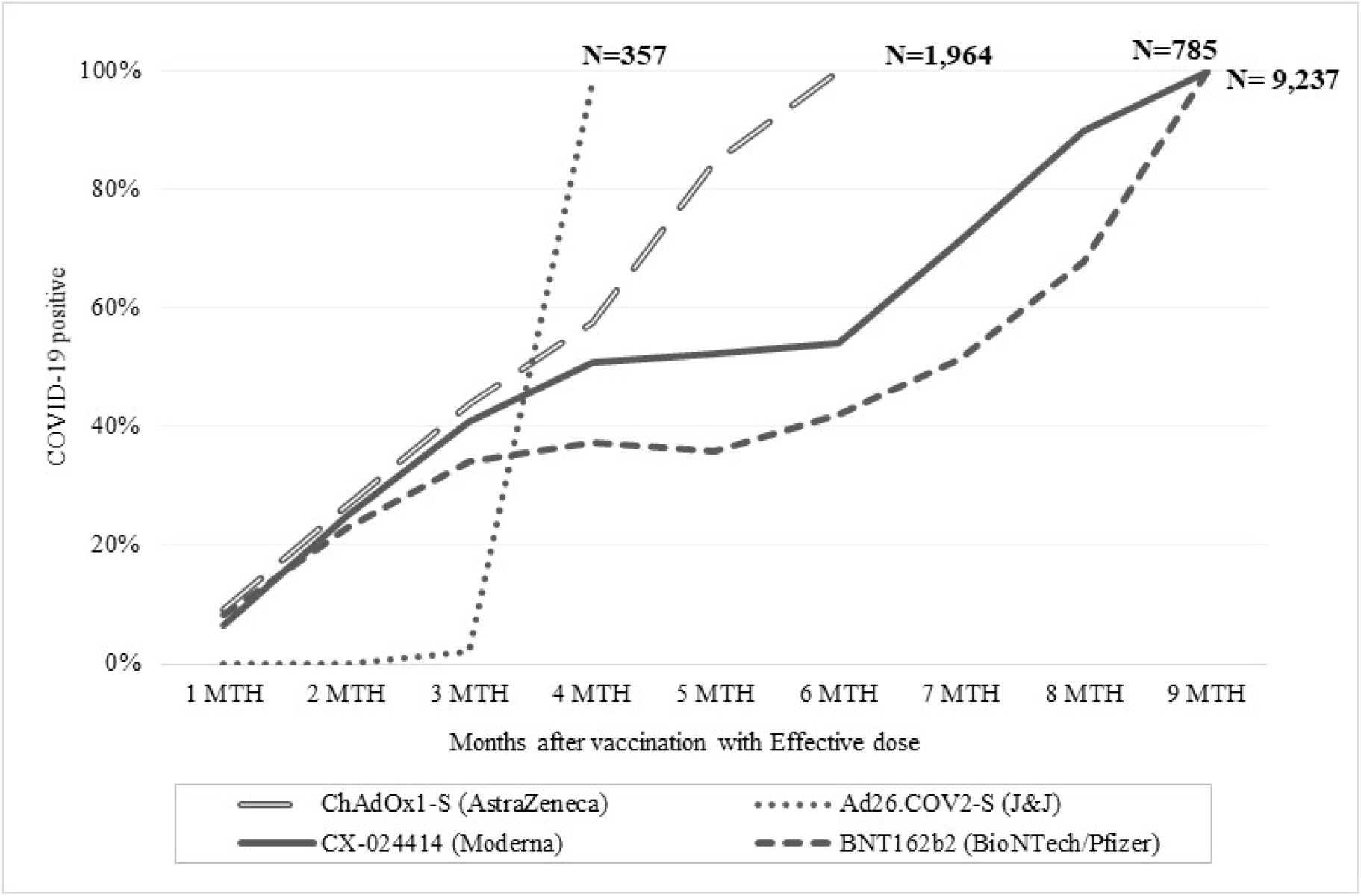
Percentage of COVID-19 positive patients stratified by number of months after vaccination with effective dose and stratified by age groups and stratified by vaccine type

On the other hand, 50% of the subjects vaccinated with ChAdOx1 nCoV-19 adenoviral (AstraZeneca) (n=1,964) became positive from the fourth month after vaccination until they were all positive by the sixth month after vaccination. 50% of the subjects vaccinated with CX-024414 mRNA (Moderna) (n=785) became positive from the fourth month after vaccination, between the fourth and sixth month the trend remained constant, and then increased to 90% in the eighth month after vaccination.

37% of the subjects vaccinated BNT162b2 mRNA vaccine (Pfizer-BioNTech) (n=9,237) became positive from the fourth month after vaccination, between the fourth and fifth month the trend remained constant, and then increased to 67% in the eighth month after vaccination.

## Discussion and Conclusion

Vaccines have been a really successful technology for controlling infectious diseases in the past. COVID19 represented a never experienced global emergency with world wide spread and high mortality rate and therefore vaccines have represented a possible solution to stop SARS-CoV-2 spreading. Indeed, since the early phase of vaccine campaign the COVID19 contagiousness have registered a decrease^5^, however few real-world studies are available on prolonged follow-up and on larger cohort population. As first consideration, according to our primary outcome, the results of this large community study based on Campania Region population (ISTAT censed citizens at December 2020 of 5.889.567) showed that vaccination with two doses of BNT162b2 or ChAdOx1 still significantly reduces the risk of new PCR-positive SARS-CoV-2 infection.

Further it is interestingly to emphasize that they also showed a strongly significant reduction of ICU hospital stay when vaccinated patients are hospitalized compared to unvaccinated subjects (Table 2). However, despite these findings, we found a rate of breakthrough infection of (7.3%) among studied population receiving an effective vaccination schedule while 0.33% of total vaccinated population in Campania Region. These showed increasing spread of positive RT-PCR according to time elapsed from second dose vaccination with highest risk after five months while only after two months for a single shot vaccine. The highest percentage of infected positive subjects were asymptomatic while the dynamic of protection varied by vaccine type, with initially similar effectiveness of both mRNA vaccines and ChAdOx1, that after 4 months become less effective with a more rapid declining of coverage for adenoviral vector-based vaccine.

Concerning Ad26.COV2-S vaccine it seems to show a waning of adequate coverage after one month after completing vaccination schedule. Those findings, however, demonstrated that vaccines are able to decrease severity of SARS-CoV-2 disease once infected. Nonetheless, it seems also clear that vaccine, even if effective to decrease severity of disease and risk of severe hospitalization, showed a decrease in their efficacy to protect against SARS-CoV-2 contagiousness throughout the time in a significant percentage but not the majority of vaccinated people. This consideration would suggest that at least in some cases, the vaccine protected against symptomatic disease but not against infection throughout the time. The possible reasons of our findings could be viral and immune related. In fact it is to underline that according to data available for Italian National Institute of Health, in Italy and therefore in Campania we have had an increase in frequency of Delta variant reaching at the end of July almost 95% of all isolated virus^16^. Thus a less effective vaccine coverage when delta variant become dominant while vaccine campaign proceed could have had a role in this breakthrough. Another explanation could be in different effects of vaccination on immunity, cellular or humoral possibly determining different infective state^17^ mainly characterized by asymptomatic subjects. These considerations would also underline the need to better understand what kind of impact asymptomatic infected vaccinated people may have on SARS-CoV-2 spreading among unvaccinated and vaccinated people too. Indeed, even if the current findings of vaccine effectiveness to protect against severe outcomes would seem to suggest that virus transmission and nasopharyngeal viral presence may have limited consequences, we could have some important consequences over time. In fact, in absence of an universal vaccination possible environments where the SARS-CoV-2 may develop elusive strategies by increasing its mutational rate or fitness could compromise vaccine efficacy. The latter event could make herd immunity less likely with possible severe evolution in particular settings of patients. Despite this could be possible future scenario, our findings may be of usefulness in preventing it. Particularly our study has two main strengths. First, we provide extensive documentation on a large cohort of breakthrough infections, based on a Regional Big Data where all COVID19 positive data are automatically evaluated, matched and analysed for their vaccine status by real time ML algorithm, minimizing error on records giving a real time scenario and trend. Second, this cohort is one of the largest presented in literature and represents all ages underwent vaccination with a very good representation of all currently approved vaccines. Therefore, in conclusion, in this study we found that although the current approved COVID19 vaccine are extremely effective in reducing hospitalization and particularly ICU, breakthrough infections occur with a breakpoint between 5^th^ and 7^th^ month after vaccination and they may carry a potential infectiveness. This event could represent a challenge, since such infections are often asymptomatic and may pose a risk to vulnerable populations. Consequently, a boost dose could be a possible strategy while awaiting the antiviral ^18, 19^ that could give us a final weapon against SARS-CoV-2. However, considering highest percentage of asymptomatic patients and that few data about their capacity to transmit SARS-CoV-2, further screening, quarantine procedure and other preventing strategies should be guaranteed in all vaccinated subjects. Finally, a surveillance approach based on the use of integrated BIG Data system to match all clinical conditions too, offer a precise and real analysis with low incidence of errors in the categorization of subjects.

## Data Availability

All data produced in the present work are contained in the manuscript

## References

1. WHO Coronavirus (COVID-19) Dashboard. https://covid19.who.int/ Accessed on 1 November 2021

2. Krammer F. SARS-CoV-2 vaccines in development. Nature. 2020 Oct;586(7830):516–527. doi: 10.1038/s41586-020-2798-3. Epub 2020 Sep 23.

3. Optimising the COVID-19 vaccination programme for maximum short-term impact. https://www.gov.uk/government/publications/prioritising-the-first-covid-19-vaccine-dose-jcvi-statement/optimising-the-covid-19-vaccination-programme-for-maximum-short-term-impact Accessed on 1 November 2021

4. Mallapaty S. Can COVID vaccines stop transmission? Scientists race to find answers. Nature. 2021 Feb 19. doi: 10.1038/d41586-021-00450-z. Epub ahead of print.

5. Impatto della vaccinazione COVID-19 sul rischio di infezione da SARS-CoV-2 e successivo ricovero e decesso in Italia https://www.epicentro.iss.it/vaccini/pdf/report-valutazione-impatto-vaccinazione-covid-19-15-mag-2021.pdf Accessed on 1 November 2021

6. Bergwerk M, Gonen T, Lustig Y, Amit S, Lipsitch M, Cohen C, Mandelboim M, Levin EG, Rubin C, Indenbaum V, Tal I, Zavitan M, Zuckerman N, Bar-Chaim A, Kreiss Y, Regev-Yochay G. Covid-19 Breakthrough Infections in Vaccinated Health Care Workers. N Engl J Med. 2021 Oct 14;385(16):1474–1484. doi: 10.1056/NEJMoa2109072. Epub 2021 Jul 28

7. Khoury J, Najjar-Debbiny R, Hanna A, Jabbour A, Abu Ahmad Y, Saffuri A, Abu-Sinni M, Shkeiri R, Elemy A, Hakim F. COVID-19 vaccine - Long term immune decline and breakthrough infections. Vaccine. 2021 Nov 26;39(48):6984–6989. doi: 10.1016/j.vaccine.2021.10.038. Epub 2021 Oct 30.

8. Pouwels KB, Pritchard E, Matthews PC, Stoesser N, Eyre DW, Vihta KD, House T, Hay J, Bell JI, Newton JN, Farrar J, Crook D, Cook D, Rourke E, Studley R, Peto TEA, Diamond I, Walker AS. Effect of Delta variant on viral burden and vaccine effectiveness against new SARS-CoV-2 infections in the UK. Nat Med. 2021 Oct 14. doi: 10.1038/s41591-021-01548-7. Epub ahead of print.

9. Perrella A, Orlando V, Trama U, Bernardi FF, Menditto E, Coscioni E. Pre-Exposure Prophylaxis with Hydroxychloroquine Does Not Prevent COVID-19 nor Virus Related Venous Thromboembolism. Viruses. 2021 Oct 13;13(10):2052. doi: 10.3390/v13102052.

10. Orlando V, Coscioni E, Guarino I, Mucherino S, Perrella A, Trama U, Limongelli G, Menditto E. Drug-utilisation profiles and COVID-19. Sci Rep. 2021 Apr 26;11(1):8913. doi: 10.1038/s41598-021-88398-y.

11. Orlando V, Rea F, Savaré L, Guarino I, Mucherino S, Perrella A, Trama U, Coscioni E, Menditto E, Corrao G. Development and validation of a clinical risk score to predict the risk of SARS-CoV-2 infection from administrative data: A population-based cohort study from Italy. PLoS One. 2021 Jan 20;16(1):e0237202. doi: 10.1371/journal.pone.0237202.

12. Bliek-Bueno K, Mucherino S, Poblador-Plou B, González-Rubio F, Aza-Pascual-Salcedo M, Orlando V, (…) Gimeno-Miguel A. Baseline Drug Treatments as Indicators of Increased Risk of COVID-19 Mortality in Spain and ItalyInt. J Environ Res Public Health. 2021 Nov 10;18(22):11786. doi: 10.3390/ijerph182211786

13. Perrella A, Carannante N, Berretta M, Rinaldi M, Maturo N, Rinaldi L. Novel Coronavirus 2019 (Sars-CoV2): a global emergency that needs new approaches? Eur Rev Med Pharmacol Sci. 2020 Feb;24(4):2162–2164.

14. Rhee C, Kanjilal S, Baker M, Klompas M. Duration of Severe Acute Respiratory Syndrome Coronavirus 2 (SARS-CoV-2) Infectivity: When Is It Safe to Discontinue Isolation? Clin Infect Dis. 2021 Apr 26;72(8):1467–1474. doi: 10.1093/cid/ciaa1249.

15. Perrella A, Brita M, Coletta F, Cotena S, De Marco G, Longobardi A, Sala C, Sannino D, Tomasello A, Perrella M, Russo G, Tarsitano M, Chetta M, Della Monica M, Orlando V, Coscioni E, Villani R. SARS-CoV-2 in Urine May Predict a Severe Evolution of COVID-19. J Clin Med. 2021 Sep 8;10(18):4061. doi: 10.3390/jcm10184061.

16. https://www.iss.it/cov19-cosa-fa-iss-varianti/asset_publisher/yJS4xO2fauqM/content/id/5807918

17. Wei J, Stoesser N, Matthews PC, Ayoubkhani D, Studley R, Bell I, Bell JI, Newton JN, Farrar J, Diamond I, Rourke E, Howarth A, Marsden BD, Hoosdally S, Jones EY, Stuart DI, Crook DW, Peto TEA, Pouwels KB, Eyre DW, Walker AS; COVID-19 Infection Survey team. Antibody responses to SARS-CoV-2 vaccines in 45,965 adults from the general population of the United Kingdom. Nat Microbiol. 2021 Sep;6(9):1140–1149. doi: 10.1038/s41564-021-00947-3. Epub 2021 Jul 21.

18. Fischer W, Eron JJ, Holman W, Cohen MS, Fang L, Szewczyk LJ, Sheahan TP, Baric R, Mollan KR, Wolfe CR, Duke ER, Azizad MM, Borroto-Esoda K, Wohl DA, Loftis AJ, Alabanza P, Lipansky F, Painter WP. Molnupiravir, an Oral Antiviral Treatment for COVID-19. medRxiv [Preprint]. 2021 Jun 17:2021.06.17.21258639. doi: 10.1101/2021.06.17.21258639.

19. Salasc F, Lahlali T, Laurent E, Rosa-Calatrava M, Pizzorno A. Treatments for COVID-19: Lessons from 2020 and new therapeutic options [published online ahead of print, 2021 Nov 18]. Curr Opin Pharmacol. 2021;doi:10.1016/j.coph.2021.11.002)

