## Supplemental data for "SARS-CoV-2 Infection Breakthrough among the non-vaccinated and vaccinated: a Real World Evidence study based on Big Data"

**SUPPLEMENTARY MATERIAL 1**

**Description of Machine Learning Process**

ML analysis was made according to python scripting model (Spyder IDE 64bit ver) to select, extract and match individuals vaccinated/negative or vaccinated/positive and to perform forecasting analysis on contagiousness and trend in coverage (ref). Obtained Data were further organized on Tableau Professional.

Briefly Machine learning algorithms take the information that represents the relationship between items in data sets and exporting them on a second software for statistical analysis or graphical representation. Further it creates models in order to forecast future trend and results. These models are nothing more than actions that will be taken by the machine to achieve a result. (Figure 1 supplementary). This approach considers three main elements: Event (e) (positive or negative Test), Category (c) (Type of Time of subject, vaccinated or unvaccinated and Time (t) according to the following formula for a trend evaluation: Time series (Ts) = (Ep/En + Cv/Cu ). Therefore data mining was automated according to a Machine Learning algorithm as in Figure 1 supplementary)

**Supplementary Figure 1** Block diagram of the machine-learning-based algorithm extraction for data mining for time series forecasting and statistical analysis. Current Paper show findings related to Output box b

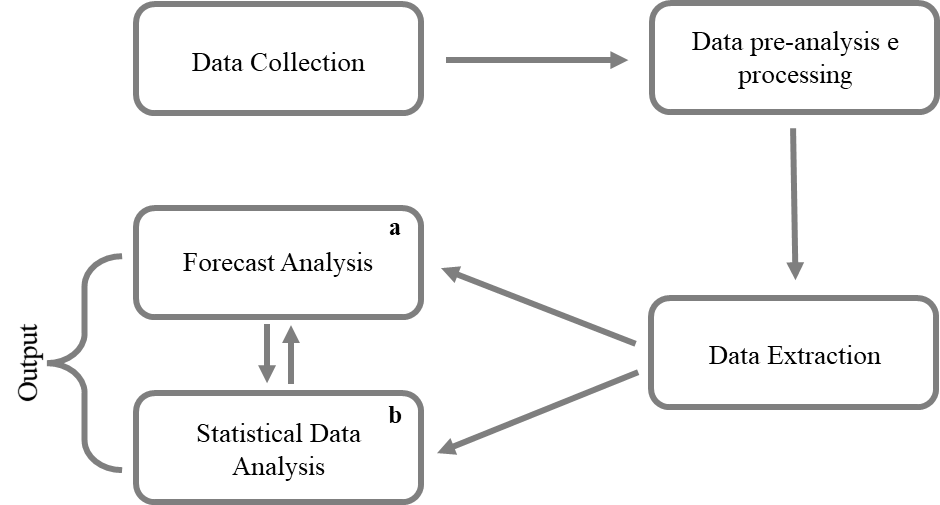
